# Waves in time, but not in space – An analysis of pandemic severity of COVID-19 in Germany based on spatio-temporal clustering

**DOI:** 10.1101/2023.01.27.23285105

**Authors:** Andreas Kuebart, Martin Stabler

**Affiliations:** Leibniz Institute for Research on Society and Space; Brandenburg Technical University; Free University of Berlin

## Abstract

While pandemic waves are often studied on the national scale, they typically are not distributed evenly within countries. This paper employs a novel approach to analyze the tempo-spatial dynamics of the COVID-19 pandemic in Germany. First, we base the analysis on a composite indicator of pandemic severity to gain a more robust understanding of the temporal dynamics of the pandemic. Second, we subdivide the pandemic during the years 2020 and 2021 into fifteen phases, each with a coherent trend of pandemic severity. Third, we analyze the patterns of spatial association during each phase. Fourth, similar types of trajectories of pandemic severity among all German counties were identified through hierarchical clustering. The results imply that the hotspots and cold spots of the first four waves of the pandemic were relatively stationary in space so that the pandemic moved in time but less in space.

## 1. Introduction

The COVID-19 pandemic, like any other pandemic, follows a pattern of acceleration and deceleration concerning case incidences among populations and regions. Over the course of a pandemic, this creates a temporal pattern in form of logistic curves often described as wave(s). These waves consist of (fast) ascents, (up to several) peaks, and declines, indicating several stages or phases of a pandemic. While pandemic waves are often analyzed on the national scale, they typically are not distributed evenly within territories (Cliff et al. 2009; Śleszyński 2021; Teller 2021; Boterman 2022; Keeler & Emch 2018). In contrast, the spatial patterns of infections and pandemic severity vary over time, which is why a tempo-spatial perspective is necessary to understand the spread of infectious diseases (Ghosh & Cartone 2021). In the case of COVID-19, several studies have found COVID-19 infections to be clustered within countries (Scarpone et al. 2020; Murgante et al. 2020; Rodríguez-Pose & Burlina 2021), and even within cities (Slijander et al. 2021). The findings on how these clusters change over time are less consistent, however. While Kim et al. (2021) find wavelike patterns in space for the case of South Korea, Boterman (2022) finds no consistent patterns for the case of the Netherlands, and D’Angelo et al. (2021) find the trajectories of Italian regions to be relatively independent of each other. This paper adds to the literature by tracing the tempo-spatial patterns of the COVID-19 pandemic in Germany throughout the years 2020 and 2021.

COVID-19 was first detected in Germany in January 2020, with four pandemic waves occurring in the following two years of the pandemic. These first two years of the COVID-19 pandemic in Germany were analyzed by developing a novel method, for which four analytical steps were performed: First, we develop a composite index of ‘pandemic severity’, which integrates the three sub-indicators of COVID-19 case incidence, the incidence of death due to COVID-19, and the incidence of COVID-19 patients on ICU (intensive care unit). Second, a phase model of fifteen pandemic phases is developed based on a change point analysis. Each of the fifteen stages in the model is coherent in terms of the trends of pandemic dynamics (e.g., rising, decreasing, stable). Third, the spatial pattern during each phase is analyzed by considering global and local spatial autocorrelation on the level of all 400 German counties. To analyze the tempo-spatial variation of pandemic severity we thus opted to analyze the spatial patterns of pandemic severity during the whole phases instead of relying on snapshots on specific dates. Fourth, hierarchical clustering was performed to identify types of similar trajectories of pandemic severity in German counties.

The paper is structured as follows: The following section describes how we proceeded and highlights methodological reasoning for the pandemic severity index, the phase model, the spatial analysis, and the cluster analysis. The subsequent section presents the findings for each step of the analysis. The final section discusses the results and concludes.

## 2. Data and methods

### 2.1. Data collection

All datasets for the pandemic severity index were accessed via Corona Daten Plattform (2022), Data on COVID-19 cases and deaths originate from the federal Robert Koch Institute (RKI), data on patients with COVID-19 on ICU were used as reported by the German Association of intensive care physicians (DIVI). The three indicators were available at the county level (Kreise and Kreisfreie Städte).

### 2.2. A composite index for pandemic severity

Studies focusing on the tempo-spatial aspects of the COVID-19 pandemic rely on the case incidence of patients who tested positive for COVID-19 almost exclusively as an indicator (Nazia et al. 2022). This is somewhat problematic, especially since the testing regime changed throughout the pandemic and potentially also varied regionally. For example, during the first wave of COVID-19 in Germany (March through May 2020), only limited capacities of PCR testing were available, whereas later PCR testing and antigen testing were widely available in 2021.

Following suggestions to combine indicators in order to add robustness to the spatio-temporal analysis of COVID-19 (Pagel & Yates 2021; Rohleder & Bozorgmehr 2022), we decided to develop a composite indicator of pandemic severity.

The pandemic severity index is composed of three indicators: the incidence of cases (IC) tested positive for COVID-19, the incidence of patients on ICU (IICU), and the incidence of registered deaths due to COVID-19 (ID, see table 1). While all three sub-indicators were gathered at the county-level, we decided to use the regional average in 96 planning regions (BBSR 2017) for IICU, because hospitals are distributed rather unevenly over German counties and especially more specialized medical infrastructure such as ICUs tend to be concentrated in larger towns and cities, which tend to be their own county.

**Table 1:** Elements of the pandemic severity composite indicator

| Indicator | Time range | Spatial resolution |
| --- | --- | --- |
| Patients tested positive for COVID-19 | 2020-02-01 -<br>2021-12-31 | 400 counties |
| Deaths of patients tested positive for COVID-19 | 2020-02-01 -<br>2021-12-31 | 400 counties |
| Patients tested positive for COVID-19 on ICU | 2020-03-04 -<br>2021-12-31 | 96 planning regions |

**Table 2:** Phases of the COVID-19 pandemic in Germany as determined through change point analysis. The trend value describes the change in pandemic severity on the national level during the respective phase.

| Phase | Duration | Length (days) | Trend |
| --- | --- | --- | --- |
| **A Prelude first wave 2020** | 2020-02-01 - 2020-03-19 | 48 | +0.057 |
| **B Surge first wave 2020** | 2020-03-20 - 2020-04-07 | 19 | +0.156 |
| **C Decline first wave 2020** | 2020-04-08 - 2020-05-25 | 48 | -0.153 |
| **D Plateau summer 2020** | 2020-05-26 - 2020-09-18 | 116 | -0.041 |
| **E Entry Surge winter wave 2020/21** | 2020-09-19 - 2020-10-10 | 22 | +0.040 |
| **F Surge winter wave 2020/21** | 2020-10-12 - 2020-11-13 | 33 | +0.333 |
| **G Further surge winter wave 2020/21** | 2020-11-14 - 2020-12-23 | 40 | +0.319 |
| **H Decline winter wave 2020/21** | 2020-12-24 - 2021-03-02 | 69 | -0.445 |
| **I Surge easter wave 2021** | 2021-03-03 - 2021-04-28 | 57 | +0.186 |
| **J Decline easter wave 2021** | 2021-04-29 - 2021-06-16 | 49 | -0.359 |
| **K Bottom summer 2021** | 2021-06-17 - 2021-07-17 | 31 | -0.061 |
| **L Entry surge summer 2021** | 2021-07-18 - 2021-08-10 | 24 | +0.019 |
| **M Entry delta wave 2021** | 2021-08-11 - 2021-10-24 | 75 | +0.186 |
| **N Surge delta wave 2021** | 2021-10-25 - 2021-12-02 | 39 | +0.462 |
| **O Decline delta wave 2021** | 2021-12-03 - 2021-12-30 | 28 | -0.201 |

The components of a composite indicator need to be scaled, weighted, and aggregated (Liborio et al. 2021). The pandemic severity index was calculated as follows: First, the rolling 14-day mean of each indicator was calculated using the zoo package in R (Zeileis & Grothendieck 2005) and they were normalized by using z-scores. Second, the scaling of the indicators was completed by minimum-maximum normalization values to a scale between 0 and 1 (Wickham & Seidel 2022). Third, the indicators were aggregated by using the arithmetic mean, in which all three indicators have the same weight. Subsequently, the composite index was rounded to four decimal places. This approach represents the common procedure to calculate composite indicators (Dialga & Giang 2017). The resulting pandemic severity index indicates the pandemic burden during a given time in each location through a dimensionless value between 0 and 1 (in practice between 0 and 0.7625). It was calculated for each day between 2020-03-01 and 2021-12-31 for each German county, resulting in 268,400 individual values. To establish a phase model on the national scale it was aggregated daily before those phases were then analyzed spatially.

### 2.3. Identifying change points of pandemic severity

A comprehensible method for defining pandemic stages is rarely found in the literature on COVID-19. Existing phase models (Ghosh & Cartone, 2020; Benita & Gasca-Sanchez 2021; Li et al. 2021; Zawbaa et al. 2022) define the beginning of each stage relatively arbitrarily based on individual indicators, such as incidence rates, mortality rates, or the implementation of countermeasures like lockdowns and social distancing. Typical types of phases include ‘beginning’, ‘outbreak’, ‘recession’, and ‘plateau’ (Li et al. 2021). Schilling et al. (2022) used a multivariant approach by combining several variables for their phase model for Germany, although their method of delineating phases remains unclear. Küchenhoff et al. (2021) calculated change points in the early course of the pandemic from March to May 2020 using a back-projection for estimated daily infections in Germany. Their retrospective exploratory analysis identified five phases for Germany within the first wave. Our phase model of the COVID-19 pandemic in Germany is based on a composite indicator and spans a longer period of several waves, during which our phases are trend coherent to serve as a heuristic for further analytical steps.

We performed a two-step change point analysis on a time series of the pandemic severity index on the national scale. A change point analysis using the binary segment approach (Scott & Knott 1974) on mean and variance was performed on the lagged difference of the pandemic severity indicator to establish rough phases, which then served as a heuristic to develop a finer model. Again, a binary segment approach was used, with a minimum segment length of 14 days. The change points were calculated using the change point package in R (Killick & Eckley 2014). The resulting phase model consists of 15 individual phases, ranging from 19 to 116 days in length.

### 2.4. Local and global autocorrelation

In a first step, the average pandemic severity in each county was calculated for each phase and mapped accordingly. Second, the global autocorrelation of pandemic severity was calculated for each day in the study period in form of Moran’s I metric (Moran 1950). Moran’s I and the related test statistic were calculated by using the sfdep package in R (Parry 2022). In a third step, each of the fifteen phases was analyzed in terms of their spatial patterns. Local indicators of spatial association (LISA) were calculated for each phase using localized Moran’s I (Anselin 1995; Sokal et al. 1998) using the sfdep package in R (Parry 2022). The contiguity matrix for the LISA analysis was established based on the k-nearest neighbor criterium with k=6 to reflect the irregular configuration of territorial borders (Ghosh & Cartone 2020), since many German counties with just one neighbor.

The LISA method has been used before to identify significant clusters of pandemic outbreaks (e.g., Scarpone et al. 2020; Siljander et al. 2022; Ghosh & Cartone 2020). However, to our knowledge, this study is the first to use this approach based on a composite indicator of pandemic severity instead of case incidence or mortality as a single indicator.

The results of a LISA analysis group the spatial units (here German counties) relative to their neighbors in local clusters of high values (high–high) or low values (low–low), and also identifies spatial outliers with (high–low) or (low–high) values. For each spatial unit, the p-value was determined through 1499 simulations. Only results with a p-value below 0.05 were considered. Since the geography of the COVID-19 pandemic is uneven in space (Scarpone et al. 2020; Rodríguez-Pose & Burlina 2021), the pandemic severity index can also help to reveal the complex tempo-spatial patterns of how the pandemic unfolded better compared to case numbers. Visualizations were created in R using the ggplot2 (Wickham 2016), and sf (Pebesma 2018) packages.

### 2.5. Hierarchical clustering of LISA sequences

To establish temporal patterns of spatial association, we further analyzed the results of the LISA analysis through hierarchical clustering, following a principle used by Bucci et al. (2022) and Mattera (2022). However, we deviate from these approaches by using the results of the LISA analysis for each county during each of the fifteen phases to calculate the dissimilarity matrix, which was used for the cluster analysis. To do so, we used Gower’s distance (Gower 1971) for the dissimilarity matrix, then hierarchical clustering was performed based on the average linkage method. Both steps were performed using the cluster package in R (Maechler et al. 2022). Only counties, which were classified into one of the four LISA categories during four or more of the fifteen phases (n=142) to avoid a sparse dissimilarity matrix. The resulting dendrogram was capped at the height of 1.0 to receive six relatively coherent clusters.

## 3. Results

### 3.1. A Phase model of the COVID-19 pandemic in Germany

Based on change point analysis of the pandemic severity index, we developed a pandemic phase model for the first two years of the pandemic. The fifteen phases of our model (table 1, chart 1) are based on trend coherence and thus differ in length. On average, each phase lasts about 47 days, although the longest phase is more than twice as long (summer plateau 2020: 116 days) and the shortest lasted only 19 days (surge of the first wave). Since the phases are based on a change point analysis using the lagged daily difference of the pandemic severity index, pandemic waves consist at least of two phases (increase and decrease). However, more complex waves can consist of several intermediate phases of acceleration and deceleration, for example, the second COVID-19 wave in Germany consists of four individual phases. Further, the periods between the phases are also considered individual phases. Among the fifteen phases during the study period, there were seven phases of increasing pandemic severity, four phases of decreasing pandemic severity, and four stable phases. This relatively fine-grained phase model serves as a heuristic to gain more specific insights into tempo-spatial patterns.

To calculate the spatial autocorrelation of pandemic severity, the Moran’s I coefficient was calculated. Spatial autocorrelation describes the association of pandemic severity among neighboring regions. The spatial autocorrelation is positive over the whole study period, which indicates a spatial association of similar values rather than a spatial association of high and low values. The Moran’s I value ranges from 0.076 in March 2020 to 0.629 during the peak of the fourth wave. This indicates the presence of spatial structure so that the distribution of pandemic severity was not random during the study period. There are notable variations of the indicator with five notable peaks in weeks 2020-w16 with 0.452, 2020-w49 with 0.484, 2021-w14 with 0.342, 2021-w33 with 0.361, and 2021-w47 with 0.629. Curiously, these peaks roughly correlate with the peaks of pandemic severity (chart 1 A and D), whereas periods with low intensity of pandemic severity tend to have lower overall spatial autocorrelation. This pattern implies that during the four waves of high pandemic severity, areas with similar pandemic severity (e.g., hot spots or cold spots) were located close to each other. Thus, we further analyzed the specific patterns of local spatial autocorrelation.

### 3.2. Local spatial association during the different phases

To analyze the distribution of pandemic severity for each of the fifteen phases we used LISA cluster maps. The LISA analysis compares each county with the level of pandemic severity in its neighboring counties. As a result, each county is associated either with high-high (HH), low-low (LL), low-high (LH), or high-low (HL) values for each of the fifteen phases and a level of significance of this value (figure 2). It should be kept in mind that the pandemic severity differed substantially between the phases.

**Figure 1:**
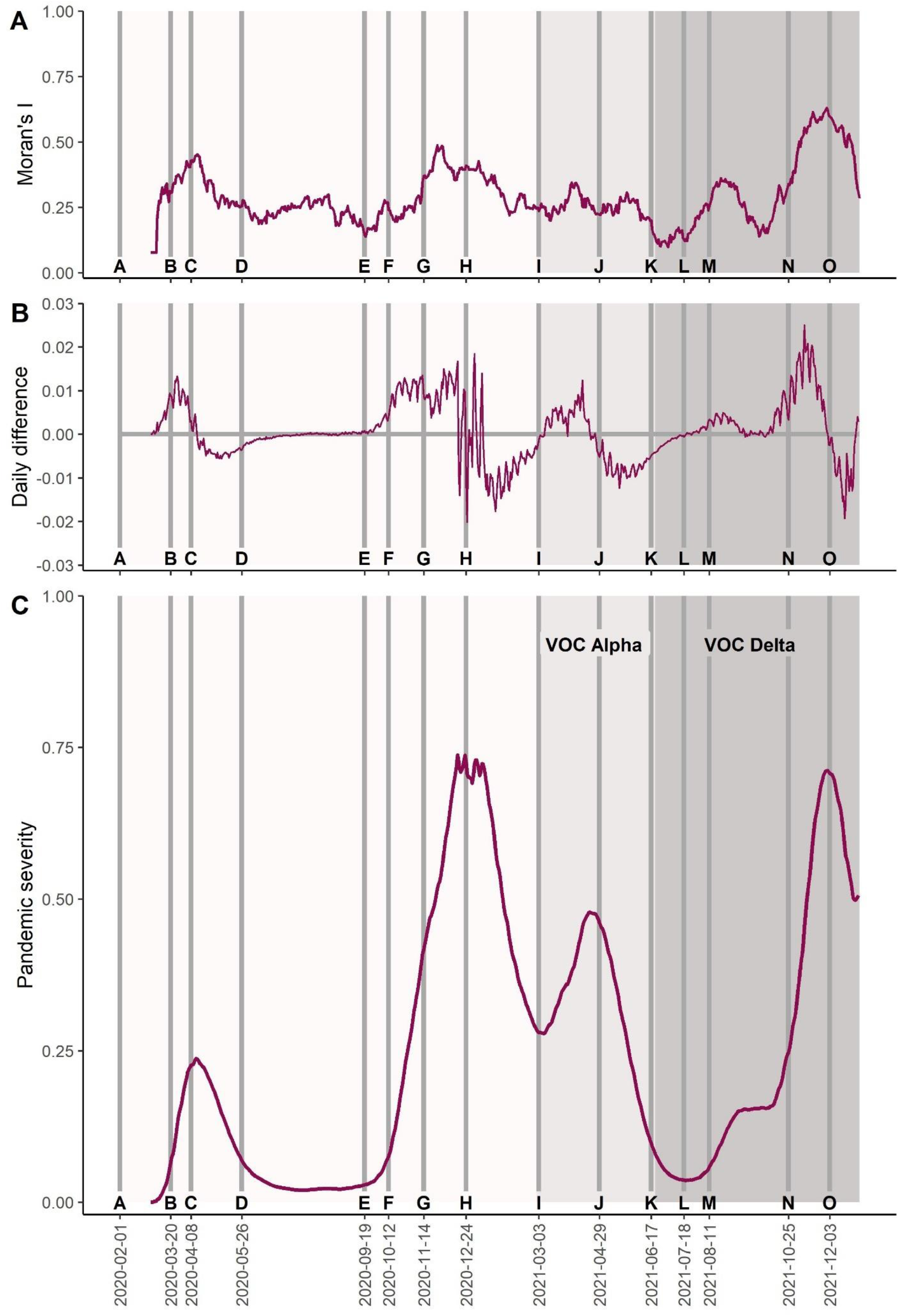
Pandemic severity in Germany and the fifteen phases of our phase model during the years 2020 and 2021(A). The phase model is based on the change points of the daily difference charts (B). The spatial autocorrelation within Germany reaches the highest levels around the peaks of the four waves (C)

**Figure 2:**
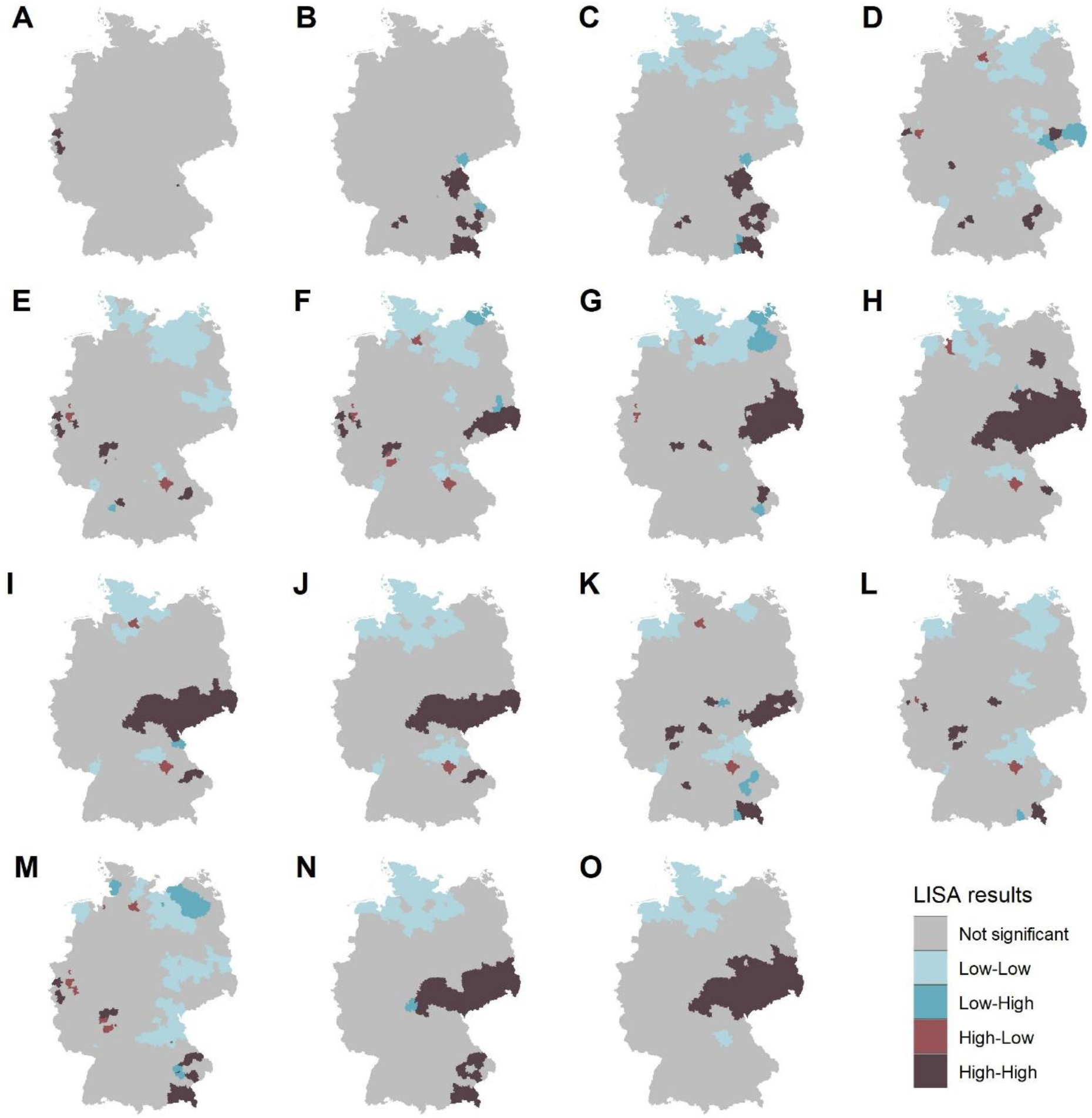
Local indicators of spatial autocorrelation of pandemic severity for the fifteen phases of the pandemic during 2020 and 2021 (A-O).

For example, a HH value during a phase of high pandemic severity (PS; such as phases B, G, or I) should mean a dire situation with many cases, deaths, and high pressure on the local healthcare system, while a HH value during a phase with modest PS should mean a less tense situation.

The resulting maps show that the pattern of local spatial autocorrelation varied substantially throughout the study period, although subsequent phases usually have similar patterns. Especially during the year 2020 (phases A-H) the overall picture changes several times, while during the year 2021, the situation seems to be more stable. Most phases are dominated by both one or more areas with several counties of HH and LL values, while HL and especially LH values are much rarer. Interestingly, the two largest counties Berlin and Hamburg both feature HL values relatively often, which indicates they are somewhat decoupled from the development around them.

While the overall pattern shows a degree of (expected) variation, some regions feature relatively stable values over time. Especially LL values are mostly concentrated in Northern Germany over much of the study period. On the other side, a concentration of HH values can be found in east-central Germany, during the phases F-K and N-O. This corresponds with a repetitive pattern during waves two, three, and four of COVID-19 in Germany (figure 2). This area, which includes much of the states of Saxony and Thuringia, and some areas in Saxony-Anhalt, and Brandenburg covers virtually all of the counties in Germany that have been hit hardest by the pandemic overall. The results of the LISA analysis imply the existence of both clusters of high and low pandemic severity, some of which seem to be relatively stable in place.

Significant values of the LISA analysis are somewhat not evenly distributed in space, since only a quarter of the possible results (fifteen phases in 400 counties) are significant and about half of the 400 counties have only a significant value for two or fewer phases, while 52% of all significant results are concentrated in 25% of the counties. Further, the proportion of significant results varies among the phases substantially between 13 of 400 counties receiving a significant result in phase A and 150 of 400 counties receiving a significant result in phase M. We further explored the tempo-spatial trajectories of counties by performing a cluster analysis on the results of the LISA analysis to identify patterns of similar trajectories of pandemic severity.

### 3.3. Types of regional trajectories

The results of the cluster analysis imply that there are several relatively stable patterns of trajectories of pandemic severities among the German counties. Six types of county trajectories were identified (figure 3), which are characterized by similar temporal patterns of pandemic severity. For the cluster analysis, we considered only counties with four or more phases with significant LISA results, so that 142 of 400 counties are included. Four types are characterized by relatively persistent values of HH (type 4, type 2, type 6) and LL (type 1), respectively, while the other types are characterized by combinations of HH and LL (type 3, type 5). Curiously, each type also has a relatively stable spatial pattern and is dominant in specific regions of Germany (figure 3B).

**Figure 3:**
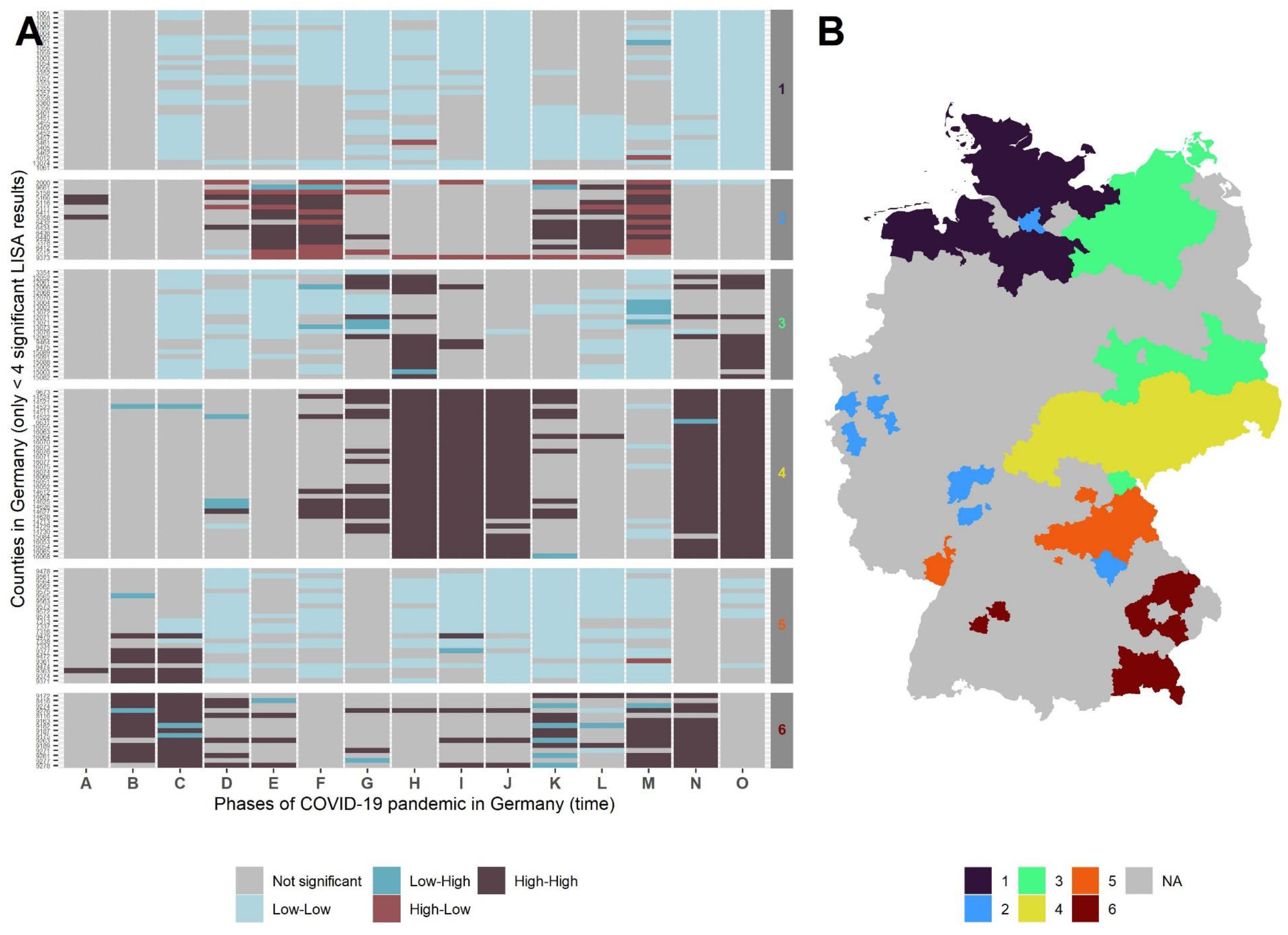
Six types of spatio-temporal trajectories of pandemic severity among German counties during the years 2020-2021. Each county with more than four significant LISA results represents a row in figure 3A and is color coded by the LISA value for each of the fifteen phases. The counties in chart 3B are color coded by the type of spatio-temporal trajectory.

The largest and most interesting type is type 4, which consists of 34 counties that are very similar in that they were classified as HH during the second (phases E-H), third (phases I-J), and fourth wave (phases M-O) of COVID-19 in Germany. However, none of the counties in this group is classified as HH during the first wave (phases A-C), which has a spatial pattern, unlike the subsequent waves. Geographically, this type consists of all counties in the state of Saxony and some states in the neighboring states of Thuringia and Saxony-Anhalt. This area includes the metropolitan areas around Dresden and Leipzig as well as rural areas.

In contrast, the second largest type 1 is characterized by 32 counties that are classified as LL throughout most of the study period. This cluster of low pandemic severity persists throughout all four waves of COVID-19 in 2020 and 2021 and is only absent during the first two phases. Geographically, this type is present in North-western Germany and includes large parts of the states of Schleswig-Holstein and Lower Saxony, as well as one county in the state of Mecklenburg-West Pomerania. While this area is predominantly rural, it also consists of parts of the metropolitan area around Hamburg, Germany’s second biggest city, albeit not Hamburg itself.

Another somewhat similar type is type 3, which is dominated by LL values during much of 2020 and in the summer of 2021. However, most counties of this type have HH values during the peak of the second wave as well as during the fourth wave in late 2021. This type is present in 22 counties in north-eastern Germany and south of Berlin in predominantly rural areas in the states of Mecklenburg-West Pomerania, Brandenburg, and Saxony-Anhalt.

Somewhat similar is type 5, which is characterized by counties that were hotspots during the first wave in early 2020 but were classified as LL for the subsequent phases. This type is present in Northern Bavaria and some parts of Rhineland-Palatinate. These 23 counties are mostly rural areas and small towns, but also encompass the Nuremberg metropolitan region. Type 5 is also characterized by counties that show HH values during the first wave. However, in contrast to the previous type, these counties also receive HH values during the fourth wave in late 2021, although the phases in between are largely insignificant. Geographically, this type is found in Southeast Bavaria and two counties in Baden-Württemberg. The 15 counties of this type are mostly rural but relatively dense areas at the edge of the Munich and Stuttgart metropolitan regions, respectively.

Finally, type 2 deviates arguably most both in terms of the temporal pattern of pandemic severity and geographically. While all other types are either patterns of HH or LL value or a combination of both, the 15 counties of type two feature dominantly HL values, which means that they deviate from their immediate surroundings. Curiously, this type is almost an inverse of type 4, so that the HL values and HH values of type 2 are concentrated not during the overall peaks of pandemic severity but right in between, during periods of lower pandemic severity. Type 2 differs geographically from the other types since it is the most dispersed of all six types. It includes mostly urban and sub-urban counties in the Rhine-Ruhr metropolitan region around Düsseldorf and in the Rhine-Main metropolitan region around Frankfurt, as well as the city of Hamburg. Generally, temporal patterns of pandemic severity align with spatial patterns in so far as the spatial patterns repeatedly occur over several waves. In the following section, these results are further discussed.

## 4. Discussion and conclusion

In this paper, we offer a comprehensive analysis of the first two years of the COVID-19 epidemic in Germany from a tempo-spatial perspective. To do so, we proceeded in four analytical steps: First, a novel composite index of pandemic severity was developed that enabled us to differentiate the dynamics of the pandemic temporally and spatially. Second, a phase model with fifteen temporally coherent phases was established through change point analysis to serve as a heuristic for spatial analysis. Third, measures of global and local autocorrelation were performed to determine regional clusters of pandemic severity for German counties.

Regions with spatial concentrations of hotspots and cold spots respectively were identified via LISA analysis. Fourth, six types of similar regional trajectories of pandemic severity were identified through hierarchical clustering among German counties. The six types of trajectories offer insights into the tempo-spatial dynamics of COVID-19 in Germany during the years 2020 and 2021. In this section, we discuss our findings by presenting empirical and methodological contributions before concluding.

Empirically, three observations are especially noteworthy from our result. First, our analysis showed that the spatial patterns of pandemic severity of COVID-19 varied substantially between the different phases of the pandemic. However, a certain pattern is visible, in that the first wave of COVID-19 in Germany (spring 2020) displays a different pattern than the subsequent three waves, while waves two, three, and four are much more similar to each other. This aligns with previous findings that the first wave was mostly driven by relocation diffusion through tourist returnees (Kuebart & Stabler 2020). The three subsequent waves emerged from a higher level of pandemic severity, which might somewhat explain the different spatial patterns. Second, it is remarkable that both hotspots and cold spots remain almost stationary from late 2020 onwards during the three most severe waves of COVID-19 in Germany. The persistent hotspot areas are almost exclusively located within an area in central Germany, including parts of the states of Saxony, Thuringia, and Saxony-Anhalt. The persistent cold spot areas on the other hand are mostly located in Northern Germany, in the states of Schleswig-Holstein, Lower Saxony, and Mecklenburg-West Pomerania. Neither hotspots nor cold spots follow patterns that would suggest obvious structural differences in terms of urbanity versus rurality or population density. While the high level of pandemic severity in Saxony has been noted before (Chilla et al. 2022), it is still remarkable how stable this area returns as an area of hotspots of pandemic severity in each of three subsequent waves. Third, it is also noteworthy from a tempo-spatial perspective that the epicenter of each wave is within this region, although waves three and four have been driven by the novel alpha and delta variants of SARS-CoV-2, respectively so that relocation diffusion should have been in an important factor. Indeed, the first hotspots of variant alpha transmission were found to be in western and northern Germany (Mitze & Rode 2022), far from the region in central Germany that would be the region with the highest concentration of pandemic severity due to variant alpha only six weeks later.

Taken together, these three empirical findings imply that the COVID-19 pandemic in Germany progressed wavelike just in temporal terms, but not spatially. This is in line with the findings of D’Angelo et al. (2021), who found the case evolution over time in Italian regions to be relatively independent of each other. As in our case, this implies that region-specific aspects trump expansion diffusion dynamics, although this does not seem to be a universal phenomenon, since Kim et al. (2021) find wavelike patterns in space for the case of South Korea and Boterman (2022) finds no consistent patterns for the case of the Netherlands. However, we conclude that for the German case with its strict non-pharmaceutical interventions and largely successful vaccination campaign during the year 2021, pandemic severity is indeed less related to relocation diffusion. Instead, regional factors such as cultural or political aspects determine the effectiveness of counter-pandemic measures and thus the local pandemic diffusion seems to be more important. Further, not just the specific regional conditions but also the timing in the pandemic process should be considered when analyzing the tempo-spatial dynamics of infectious diseases.

Methodologically, also three conclusions can be drawn from the approach presented in this paper. First, the tempo-spatial clustering of time series of pandemic severity is a valuable tool to explore the dynamics of infectious outbreaks. While this approach has been successfully applied before (Bucci et al. 2022; Mattera 2022), we combined this approach with LISA analysis (Anselin 1995). In our opinion, this has the advantage that only those regions that deviate significantly from the national average are considered (e.g., hotspots and cold spots of pandemic severity), which allows focusing on the most relevant processes or regions for each phase of the pandemic. Second, a phase model based on change point analysis was a valuable heuristic to further analyze spatio-temporal dynamics of COVID-19. The choice of which points in time or periods to compare is crucial for tempo-spatial analysis. Most studies either use relatively arbitrary units such as weekly or bi-weekly (Siljander et al. 2022) intervals. In contrast, we decided to develop a phase model that distinguished different levels of pandemic severity, which proved valuable to contextualize spatial differences. Third, the use of a composite indicator as the base for subsequent analytical steps was useful to add robustness to the analysis. To follow the calls for combining different indicators when analyzing the spread of COVID-19 (Pagel & Yates 2021; Rohleder & Bozorgmehr 2022), we developed an index of pandemic severity that incorporated the incidence of patients tested positive for COVID-19, the incidence of patients with COVID-19 on ICU, and the incidence of registered deaths due to COVID-19. Although the timespan analyzed in this paper included only 21 months, the conditions under which data were collected changed drastically in several regards. Factors that varied over time included the testing regime, the chain of reporting itself, new variants of COVID-19, and increasing levels of immunity within the population due to infections and vaccinations. Therefore, we argue that combining indicators enhances comparability over longer timespans and adds robustness to the analysis. However, this is certainly not limited to the sub-indicators used here. Other indicators could and should be included in future attempts, for example, data originating from wastewater monitoring.

Some limitations of the research presented here should be considered. First, we did not include demographic factors in our pandemic severity index. Spatial variation in factors such as age distribution or prevalence of chronic diseases could influence the spatial distribution of deaths by COVID-19 and would thus somewhat influence the spatial pattern of pandemic severity per our index. Relatedly, data that describes the conditions of local infections would be helpful to contextualize local pandemic severity. Especially regional data on outbreaks and imported infections would be helpful but were not made available by the German authorities. Second, the use of data that is only reported for territorial administrative areas presents a limitation, since it might obscure patterns on a smaller scale, for example in border regions (Scarpone et al. 2020, Chilla et al. 2022). On the other hand, an analysis of spatial patterns on an even larger scale (e.g., NUTS 2 regions) might be suitable to reduce noise in the cluster analysis. In conclusion, both more fine-grained analysis of hotspot regions and analysis on a higher spatial order can be helpful steps to gain a better understanding of the dynamics of the pandemic.

In conclusion, the spatio-temporal dynamics of COVID-19 offer fascinating insights into how pathogens spread in contemporary societies. While it would be misleading to put too much emphasis on the territorial dimension of space in the era of “post-Westphalian pathogens” (Fidler 2003), the “territorial immune system” of non-pharmaceutical interventions visible during the COVID-19 pandemic has proven the relevance of territories and their impact on the pandemic process.

## Data Availability

All data produced in the present study are available upon reasonable request to the authors

## References

Anselin L. Local Indicators of Spatial Association—LISA. Geogr Anal 1995;27:93–115. https://doi.org/10.1111/j.1538-4632.1995.tb00338.x.

BBSR (2017). Laufende Raumbeobachtung – Raumabgrenzungen. Bundesinstitut für Bau-, Stadt-und Raumforschung. https://www.bbsr.bund.de/BBSR/DE/forschung/ raumbeobachtung/Raumabgrenzungen/deutschland/regionen/Raumordnungsregionen/ raumordnungsregionen.html. Checked 25.12.2022

Benita F, Gasca-Sanchez F. The main factors influencing COVID-19 spread and deaths in Mexico: A comparison between phases I and II. Appl Geogr 2021;134:102523. https://doi.org/10.1016/j.apgeog.2021.102523.

Boterman W. Population density and SARS-CoV-2 pandemic: Comparing the geography of different waves in the Netherlands. Urban Stud 2022:004209802210871. https://doi.org/10.1177/00420980221087165.

Bucci A, Ippoliti L, Valentini P, Fontanella S. Clustering spatio-temporal series of confirmed COVID-19 deaths in Europe. Spat Stat 2022;49:100543. https://doi.org/10.1016/j.spasta.2021.100543.

Chilla T, Große T, Hippe S, Walker BB. COVID-19 incidence in border regions: spatiotemporal patterns and border control measures. Public Health 2022;202:80–3. https://doi.org/10.1016/j.puhe.2021.11.006.

Cliff AD, Smallman-Raynor M, Haggett P, Stroup D, Thacker S. Infectious Diseases: A Geographical Analysis: Emergence and Re-emergence. Oxford: Oxford University Press; 2009.

D’angelo N, Abbruzzo A, Adelfio G. Spatio-temporal spread pattern of covid-19 in italy. Mathematics 2021;9. https://doi.org/10.3390/math9192454.

Dialga I, Thi Hang Giang L. Highlighting Methodological Limitations in the Steps of Composite Indicators Construction. Soc Indic Res 2017;131:441–65. https://doi.org/10.1007/s11205-016-1263-z.

Fidler DP. SARS: Political Pathology of the First Post-Westphalian Pathogen. J Law, Med Ethics 2003;31:485–505.

Ghosh P, Cartone A. A Spatio-temporal analysis of COVID-19 outbreak in Italy. Reg Sci Policy Pract 2020;12:1047–62. https://doi.org/10.1111/rsp3.12376.

Gower JC. A General Coefficient of Similarity and Some of Its Properties. vol. 27. 1971.

Keeler C, Emch M. Infectious-disease geography. Routledge Handb. Heal. Geogr., Routledge; 2018, p. 45–51. https://doi.org/10.4324/9781315104584-7.

Killick R, Eckley IA. {changepoint}: An {R} Package for Changepoint Analysis. J Stat Softw 2014;58:1–19.

Kim S, Kim M, Lee S, Lee YJ. Discovering spatiotemporal patterns of COVID-19 pandemic in South Korea. Sci Rep 2021;11. https://doi.org/10.1038/s41598-021-03487-2.

Küchenhoff H, Günther F, Höhle M, Bender A. Analysis of the early COVID-19 epidemic curve in Germany by regression models with change points. Epidemiol Infect 2021. https://doi.org/10.1017/S0950268821000558.

Kuebart A, Stabler M. Infectious Diseases as Socio-Spatial Processes: The COVID-19 Outbreak In Germany. Tijdschr Voor Econ En Soc Geogr 2020;111:482–96. https://doi.org/10.1111/tesg.12429.

Li M, Guo X, Wang X. Retrospective prediction of the epidemic trend of COVID-19 in Wuhan at four phases. J Med Virol 2021;93:2493–8. https://doi.org/10.1002/jmv.26781.

Libório MP, Ekel PY, de Abreu JF, Laudares S. Factors that most expose countries to COVID-19: a composite indicators-based approach. GeoJournal 2022;87:5435–49. https://doi.org/10.1007/s10708-021-10557-5.

Mattera R. A weighted approach for spatio-temporal clustering of COVID-19 spread in Italy. Spat Spatiotemporal Epidemiol 2022;41. https://doi.org/10.1016/J.SSTE.2022.100500.

Mitze T, Rode J. Early-stage spatial disease surveillance of novel SARS-CoV-2 variants of concern in Germany with crowdsourced data. Sci Rep 2022;12. https://doi.org/10.1038/s41598-021-04573-1.

Moran P. Notes on continous stochastic phenomena. Biometrika 1950;37:17–23. https://doi.org/10.1093/biomet/37.1-2.17.

Murgante B, Borruso G, Balletto G, Castiglia P, Dettori M. Why Italy first? Health, geographical and planning aspects of the COVID-19 outbreak. Sustain 2020;12. https://doi.org/10.3390/su12125064.

Nazia N, Butt ZA, Bedard ML, Tang W-C, Sehar H, Law J. Methods Used in the Spatial and Spatiotemporal Analysis of COVID-19 Epidemiology: A Systematic Review. Int J Environ Res Public Health 2022;19:8267. https://doi.org/10.3390/ijerph19148267.

Pagel C, Yates C. Tackling the panddemic with (biased) data. Science (80-) 2021.Parry J. sfdep: Spatial Dependence for Simple Features 2022.

Parry, Josiah. 2022. “Sfdep: Spatial Dependence for Simple Features.” cran.r-project.org/package=sfdep.

Pebesma E. Simple Features for R: Standardized Support for Spatial Vector Data. R J 2018;10:439–46. https://doi.org/10.32614/RJ-2018-009.

Rodríguez-Pose A, Burlina C. Institutions and the uneven geography of the first wave of the COVID-19 pandemic. J Reg Sci 2021;61:728–52. https://doi.org/10.1111/jors.12541.

Rohleder S, Bozorgmehr K. Monitoring the spatiotemporal epidemiology of Covid-19 incidence and mortality: A small-area analysis in Germany. Spat Spatiotemporal Epidemiol 2021;38. https://doi.org/10.1016/j.sste.2021.100433.

Scarpone C, Brinkmann ST, Große T, Sonnenwald D, Fuchs M, Walker BB. A multimethod approach for county-scale geospatial analysis of emerging infectious diseases: A cross-sectional case study of COVID-19 incidence in Germany. Int J Health Geogr 2020;19. https://doi.org/10.1186/s12942-020-00225-1.

Schilling J, Buda S, Tolksdorf K. Zweite Aktualisierung der „Retrospektiven Phaseneinteilung der COVID-19-Pandemie in Deutschland”. Epididemiologisches Bull 2022;10:3–5. https://doi.org/10.25646/9787.

Scott AJ, Knott M. A Cluster Analysis Method for Grouping Means in the Analysis of Variance. Biometrics 1974;30:507. https://doi.org/10.2307/2529204.

Siljander M, Uusitalo R, Pellikka P, Isosomppi S, Vapalahti O. Spatiotemporal clustering patterns and sociodemographic determinants of COVID-19 (SARS-CoV-2) infections in Helsinki, Finland. Spat Spatiotemporal Epidemiol 2022;41. https://doi.org/10.1016/j.sste.2022.100493.

Śleszyński P, Blaszke M. THE CITY CHALLENGES AND EXTERNAL AGENTS. METHODS, TOOLS AND BEST PRACTICES 3 (2020). J L Use, Mobil Environ 2020. Sokal RR, Oden NL, Thomson BA. Local spatial autocorrelation in a biological model. Geogr Anal 1998;30:331–54. https://doi.org/10.1111/j.1538-4632.1998.tb00406.x.

Teller J. Urban density and Covid-19: towards an adaptive approach. Build Cities 2021;2:150–65. https://doi.org/10.5334/bc.89.

Wickham H. ggplot2: Elegant Graphics for Data Analysis. Springer-Verlag New York; 2016.

Wickham H, Seidel D (2022). _scales: Scale Functions for Visualization_. R package version 1.2.0, https://CRAN.R-project.org/package=scales.

Zawbaa HM, Osama H, El-Gendy A, Saeed H, Harb HS, Madney YM, et al. Effect of mutation and vaccination on spread, severity, and mortality of COVID-19 disease. J Med Virol 2022;94:197–204. https://doi.org/10.1002/jmv.27293.

Zeileis A, Grothendieck G. zoo: S3 Infrastructure for Regular and Irregular Time Series. J Stat Softw 2005;14:1–27. https://doi.org/10.18637/jss.v014.i06.

